# Heterogeneous Effects of Sodium-Glucose Cotransporter-2 Inhibitors on Acute Kidney Injury: A Causal Learning Approach

**DOI:** 10.1101/2025.11.23.25340831

**Authors:** Hao Dai, Yao An Lee, Jiang Bian, Jingchuan Guo

**Affiliations:** Department of Biostatistics & Health Data Science, Indiana University, Indianapolis, IN, USA; Department of Pharmaceutical Outcomes & Policy, University of Florida, Gainesville, FL, USA; Center for Biomedical Informatics, Regenstrief Institute, Indianapolis, IN, USA; Weldon School of Biomedical Engineering, Purdue University, IN, USA; Indiana University Melvin and Bren Simon Comprehensive Cancer Center, Indianapolis, IN, USA; Department of Pharmacy Practice, Purdue University College of Pharmacy

**Keywords:** Heterogenous treatment effect, Sodium–glucose cotransporter-2 inhibitors, Acute kidney injury, Target trial emulation, Causal Pathways

## Abstract

**Background:** Sodium–glucose cotransporter-2 inhibitors (SGLT2is) have been associated with lower risk of acute kidney injury (AKI), but existing studies rarely explore heterogeneous treatment effects or underlying causal pathways. We applied a comprehensive causal-learning framework to evaluate both overall and subgroup-specific effects of SGLT2i therapy on AKI.

**Methods:** Using a new-user, active-comparator target trial emulation in the OneFlorida+ data (2014–2023), we estimated individualized and average treatment effects with a doubly robust meta-learner, assessed heterogeneity via subgroup and decision-tree analyses, and used causal structure learning and mediation methods to identify mechanistic pathways linking treatment to AKI.

**Results:** SGLT2 inhibitors were associated with a significant reduction in AKI compared with other second-line glucose-lowering drugs, with an average individual treatment effect of **−0.0039** (95% CI: −0.0065 to −0.0014). Kaplan-Meier curves demonstrated consistently lower cumulative AKI incidence among SGLT2i users. Subgroup analyses revealed substantial heterogeneity: protective effects were strongest in younger adults, males, and non-Hispanic Black patients, whereas benefits were attenuated in older adults, females, and those with baseline CKD. Decision tree-based heterogeneity modeling further identified atrial fibrillation, anemia, and antiparkinson agent use as key effect modifiers.

Causal structure learning highlighted atrial fibrillation, anemia, chronic kidney disease, and eGFR as central intermediating nodes. Mediation analyses showed that most of the benefit operated through direct pathways (ADE ≈ −0.0034 to −0.0035), while anemia **and heart failure** contributed small but statistically significant indirect effects.

**Conclusion:** SGLT2 inhibitors reduce AKI risk, but effects vary meaningfully across clinical subgroups and are partially mediated through interconnected cardio-renal pathways. Causal-learning methods provide mechanistic insight beyond average associations and may support more individualized SGLT2i therapy for AKI prevention.

## Introduction

Acute kidney injury (AKI) is a frequent and serious complication among individuals with type 2 diabetes (T2D), contributing substantially to morbidity, mortality, and healthcare burden.^1^ As diabetes prevalence continues to rise globally, identifying therapies that can mitigate kidney-related complications has become a major clinical and public health priority. In recent years, sodium–glucose cotransporter-2 inhibitors (SGLT2is) have emerged as an important class of glucose-lowering agents with benefits extending beyond glycemic control. Large randomized trials and real-world studies have consistently demonstrated the nephroprotective effects of SGLT2is, including reductions in albuminuria, slowing of chronic kidney disease (CKD) progression, and decreased rates of kidney failure.^2,3^ These findings have led to their widespread adoption and endorsement in clinical guidelines for comprehensive diabetes management.^4^

Among the diverse renal outcomes examined, a growing body of evidence suggests that SGLT2i therapy may also reduce the risk of acute kidney injury. Although AKI was initially a safety concern for SGLT2is due to osmotic diuresis and hemodynamic effects, subsequent studies have reported neutral or protective associations, indicating a lower incidence of AKI among SGLT2i users compared with other glucose-lowering drugs.^5–7^ Despite encouraging evidence, important uncertainties remain. Randomized trials, though methodologically rigorous, are typically underpowered to detect AKI events, which are relatively rare, and often lack the population diversity observed in routine clinical care. Observational studies have similarly reported a protective association between SGLT2i use and reduced AKI risk, but most have relied on exploratory, regression-based analyses that estimate only average associations. These conventional approaches seldom investigate whether the protective effect varies across patient subgroups, leaving heterogeneous treatment effects largely unknown. Likewise, the underlying mechanisms remain poorly characterized, as few studies have examined causal pathways or identified intermediate clinical factors that may mediate SGLT2i-related kidney benefits. Consequently, neither randomized evidence nor existing real-world analyses provides clear insight into who benefits most from SGLT2i therapy or how these renal benefits arise. These gaps highlight the need for a comprehensive causal-learning framework capable of estimating individualized treatment effects, uncovering causal pathways, and empirically validating mediators—advancing the field beyond simple association toward a mechanistic and patient-specific understanding of how SGLT2 inhibitors influence AKI risk.

Causal machine-learning approaches,^8^ such as doubly robust meta-learners^9^ for estimating heterogeneous treatment effects and data-driven causal structure learning for uncovering causal pathways, provide a rigorous methodological framework to strengthen causal inference in observational pharmacoepidemiology. These tools allow for flexible model specification, rigorous orthogonalization to mitigate overfitting bias, and improved exploration of effect heterogeneity across diverse patient subgroups. Furthermore, causal structure learning and mediation analyses can reveal underlying mechanisms through which SGLT2is may influence AKI risk, providing insights that extend beyond average treatment effects.

To address existing evidence gaps and methodological limitations, we aimed to evaluate the overall association between SGLT2i use and the risk of AKI using a comprehensive causal learning framework. Leveraging the large, diverse, and longitudinal OneFlorida+ Data Trust, we combined target trial emulation with advanced causal machine learning to estimate average and individualized treatment effects, identify heterogeneous risk profiles, and explore potential causal pathways. Through this multilayered analytical strategy, our study seeks to generate rigorous, mechanism-informed evidence regarding the renal safety profile of SGLT2 inhibitors in routine clinical practice.

## Methods

### Data source

Data were drawn from the OneFlorida+ Data Trust, a centralized repository maintained by the OneFlorida+ Clinical Research Consortium.^10^ OneFlorida+ links longitudinal EHRs with ancillary sources (e.g., Medicaid/Medicare claims, death records, and tumor registries) and includes demographics, diagnoses, medications, procedures, vital signs, and laboratories across participating systems.^11^ Source data covered January 1, 2012–June 30, 2023; cohort spanned January 1, 2014–June 30, 2023.

### Study design

This study employed a retrospective cohort design with a new-user approach. An intention-to-treat analysis was conducted to compare sodium–glucose cotransporter-2 inhibitors (SGLT2is) with other second-line glucose-lowering drugs (GLDs)—sulfonylureas, thiazolidinediones, dipeptidyl peptidase-4 inhibitors (DPP-4i), α-glucosidase inhibitors, and meglitinides—on the risk of acute kidney injury (AKI) among adults with type 2 diabetes (T2D). Exposure definitions are summarized in **Table S1,** and the overall design is illustrated in **Figure 1**. The study was approved by the University of Florida Institutional Review Board (IRB202201196).

**Figure 1.**
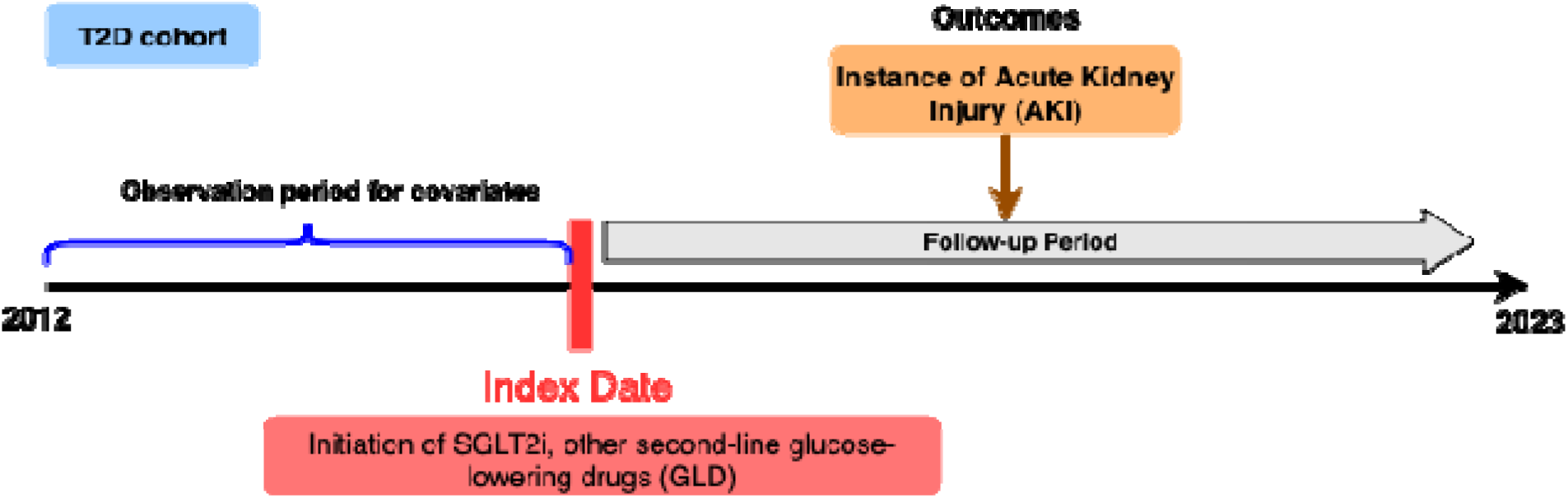
Overview of study design: new users of a SGLT2i, or other second-line glucose-lowering drug (GLD). Other second-line GLDs include sulfonylureas, thiazolidinediones, DPP4i, α-glucosidase inhibitors, or meglitinides

### Eligibility criteria

We included people who initiated treatment with a SGLT2i or other second-line GLDs in OneFlorida+ between January 1, 2014 and June 30, 2023. Other second-line GLDs were selected as the comparator group to help reduce confounding by indication (**Table S1**).^4^ The index date was the first recorded prescription for an SGLT2 inhibitor or an active-comparator GLD without a previous prescription for either drug in the previous year. We excluded individuals who were younger than 18 years at index, had a diagnosis of type 1 diabetes, gestational diabetes, or end-stage renal disease (ESRD) on or before index, or had no clinical encounters prior to index. Operational definitions and code lists are provided in **Table S2**.

### Outcome and follow-up

The primary endpoint was incident AKI during follow-up, identified via ICD-9/ICD-10 diagnosis codes (**Table S2**). We used an intention-to-treat approach, analyzing participants according to their initial treatment regardless of subsequent changes. Patients’ follow-up began at the index date and continued until the earliest occurrence of one of the following censoring events: the outcome of interest, death, loss to follow-up (the date of the last recorded clinical encounter), or the end of the study period.

### Baseline covariates

Baseline covariates encompassed a range of demographic, clinical, and pharmacological characteristics, as detailed in **Table 1**. Demographic variables included age, sex, race/ethnicity, smoking status, and insurance type. Clinical covariates comprised comorbid conditions (e.g., cardiovascular disease, cerebrovascular disease and neuropathy), clinical observations (e.g., BMI, blood pressure), and medication (e.g., insulin, opioids, and statins).

**Table 1.**
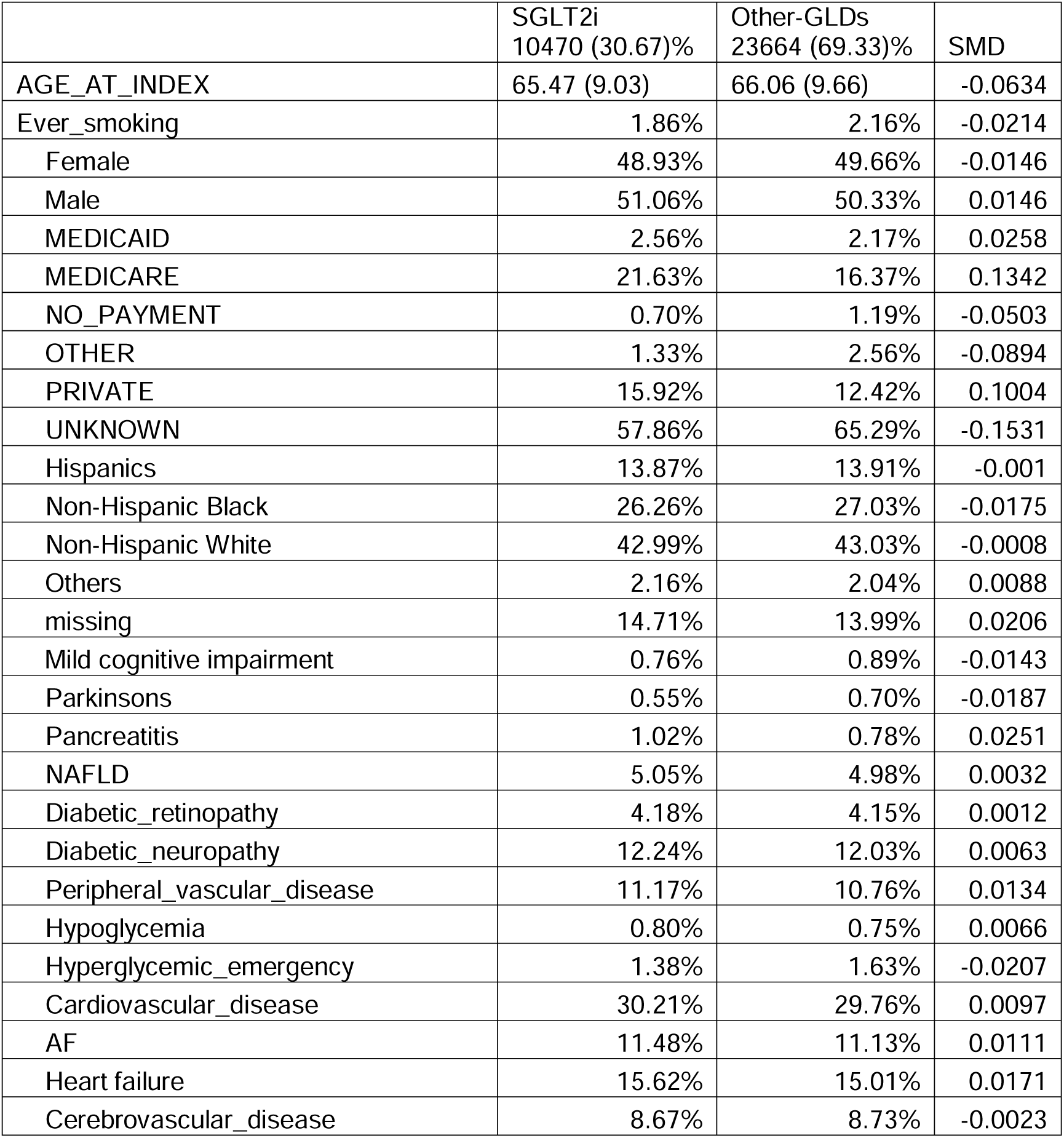
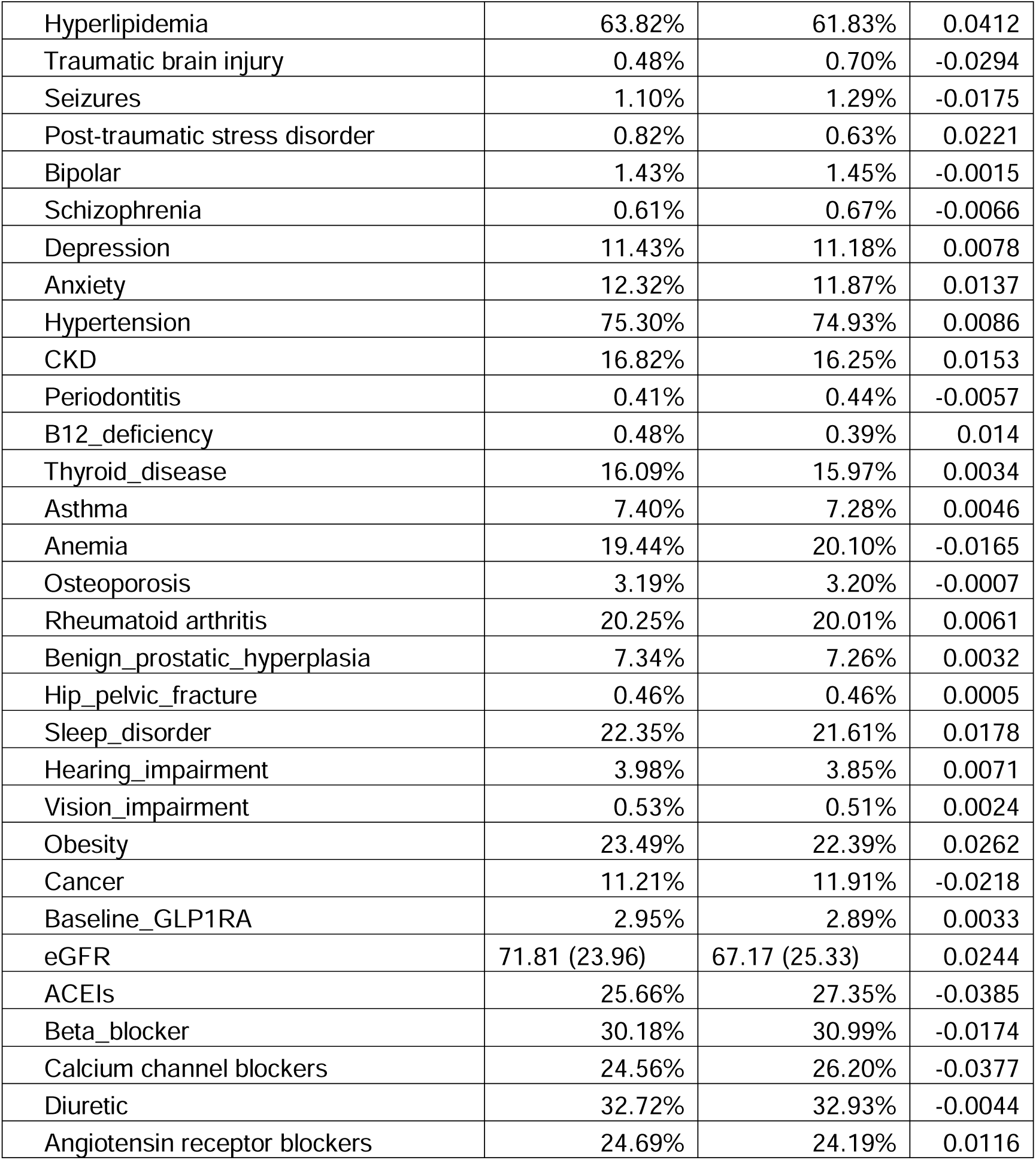
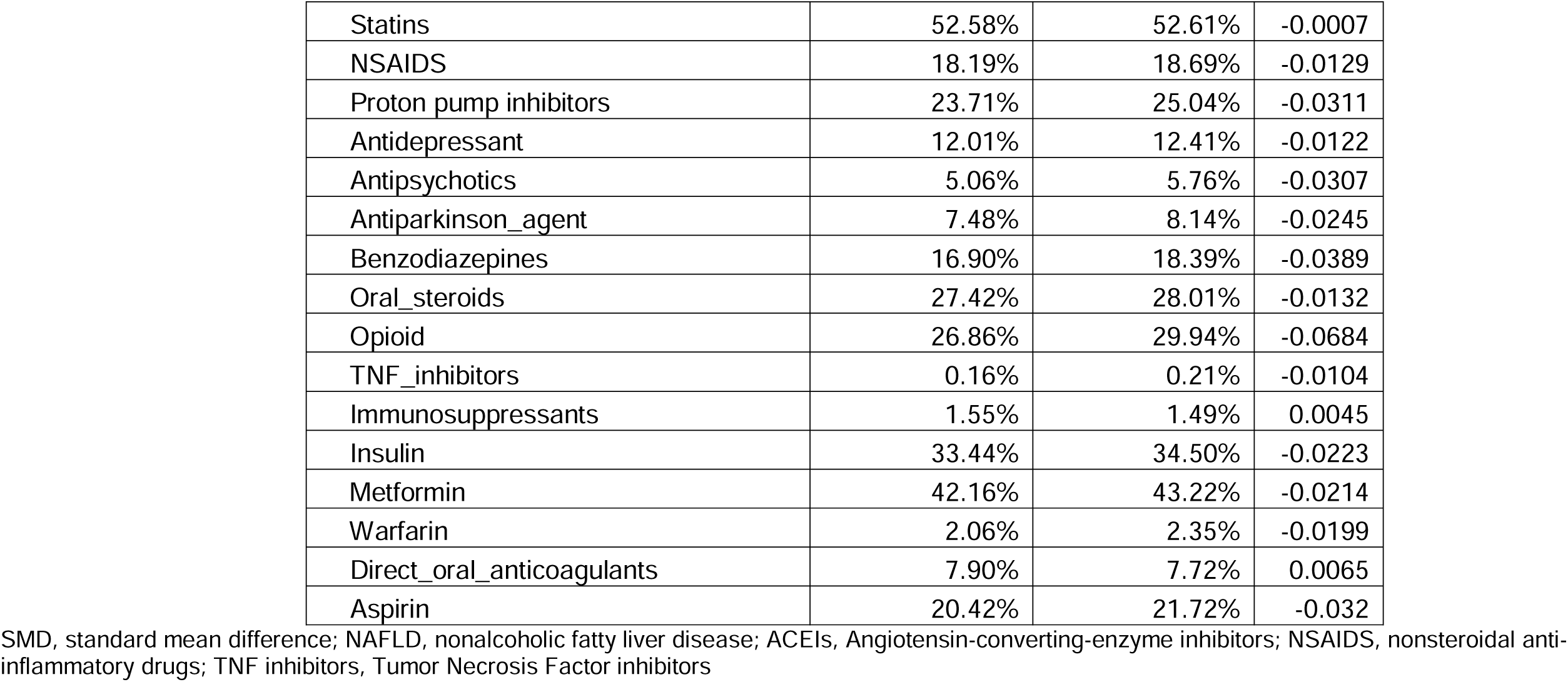
Weighted characteristics.

We structured the causal analysis into three main steps: (1) ***Primary effect estimation*** through target trial emulation, (2) heterogeneous treatment effect estimation, and (3) causal discovery, path decomposition, and mediation analysis. As shown in **Figure 2**.

**Figure 2.**
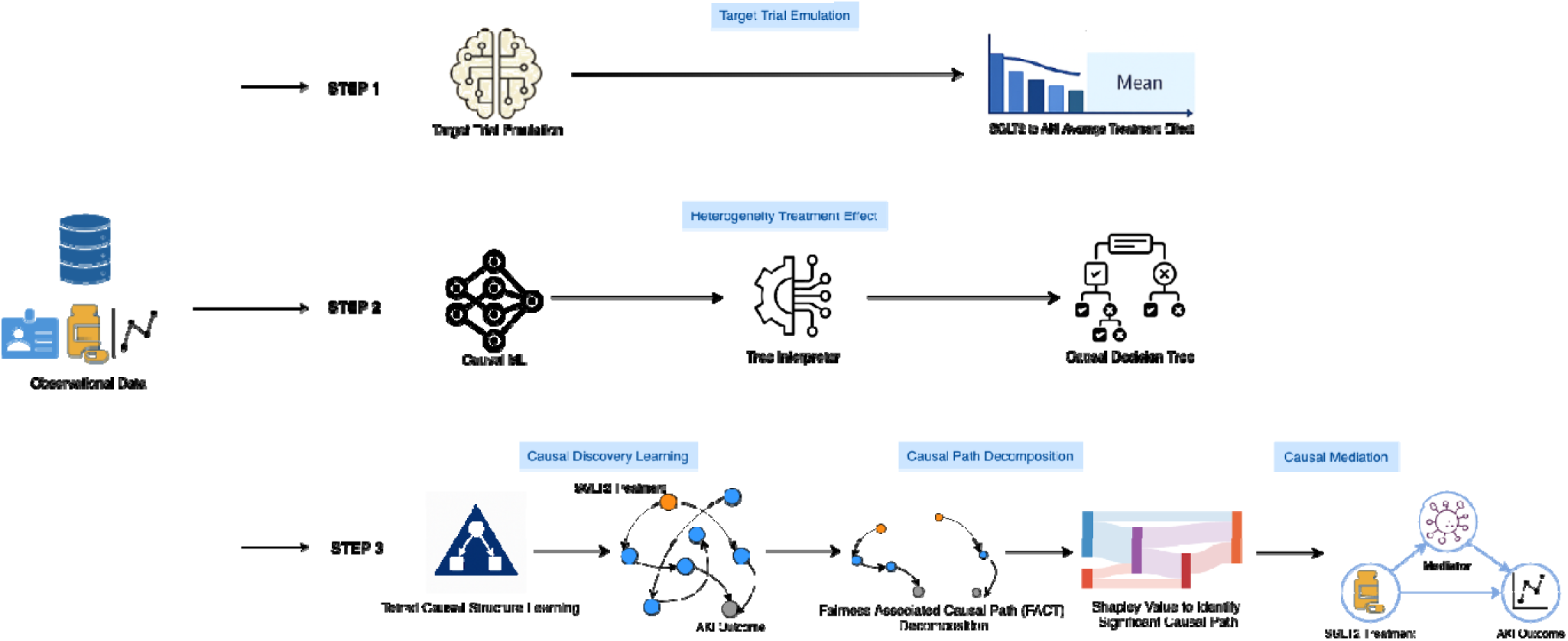
Overview of causal learning analysis

### Step 1: Primary effect estimation

We emulated the trial using a new-user, active-comparator design. Treatment assignment was modeled with a propensity score estimated from baseline covariates using a logistic regression learner with standardized inputs and hyperparameters optimized via five-fold cross-validation (GridSearchCV). The clinical outcome model was fitted using a Lasso regression with standardized predictors, and its penalty parameters were similarly tuned by cross-validated grid search to minimize mean squared error. Both nuisance models, the propensity score and outcome regression, were then incorporated into a doubly-robust meta-learner,^9^ which applied orthogonalization and five-fold cross-fitting to remove overfitting bias and obtain individualized treatment effect (ITE) estimates for each patient. The dataset was randomly split into training (70%) and testing (30%) subsets; all model fitting and hyperparameter selection were performed on the training data to prevent information leakage, and the held-out test set was used to evaluate model generalization and covariate balance.^12^ ITEs were aggregated to population-level effects using inverse probability of treatment weighting (IPTW) with stabilized, truncated weights and robust variance estimation.^13^ Time-to-event models (weighted Cox proportional hazards) yielded hazard ratios (HRs) and 95% CIs for AKI under the intention-to-treat contrast, quantifying the relative risk of kidney injury associated with SGLT2i initiation compared with the comparator therapies. Kaplan–Meier curves were generated to illustrate the cumulative probability of AKI over time between treatment groups, providing an intuitive visualization of risk divergence throughout follow-up. The proportional-hazards assumption was assessed to ensure model validity, and prespecified subgroup analyses (age, sex, race/ethnicity, baseline CKD) were performed to evaluate treatment effect heterogeneity through interaction testing and stratified modeling.

### Step 2: Heterogeneous treatment effects

To probe heterogeneity, we explored heterogeneous treatment effects (HTEs) using the ITE estimates obtained from the DML framework in Step 1. To identify clinically meaningful subgroups with differential responses to SGLT2i therapy, we applied a decision tree algorithm that recursively partitioned patients based on baseline characteristics to maximize between-group variation in treatment benefit or risk.^14^ This approach generated an interpretable stratification of the study population, highlighting key effect modifiers that contribute to heterogeneity. We subsequently conducted subgroup analyses within these tree-derived strata to examine time-to-event outcomes and assess the consistency and magnitude of treatment differences across data-driven subgroups, thereby validating and contextualizing the model-identified heterogeneity in treatment effects.

### Step 3: Causal discovery, path decomposition, and mediation

We applied a data-driven causal discovery method, Best Order Score Search (BOSS),^15^ to uncover the underlying directional relationships among demographic, clinical, and treatment variables. Using this approach, we estimated the underlying causal graph and identified potential mediating and confounding pathways that may shape treatment response. Building upon the learned graph, we employed the Fairness-Aware Causal Path Decomposition (FACTS) framework^16^ to decompose the overall treatment effect into distinct causal pathways, explicitly accounting for fairness-related factors to ensure that model-derived inferences were not driven by socially sensitive variables. Finally, we conducted causal mediation analyses to quantify the extent to which intermediate clinical conditions (e.g., atrial fibrillation, anemia, or heart failure status) mediated the total treatment effect, thereby providing mechanistic insights into how SGLT2i therapy may influence acute kidney injury risk through specific causal channels.

## Results

### Cohort Characteristics

A total of 34,134 adults with type 2 diabetes initiated either an SGLT2 inhibitor (n = 10,470; 30.67%) or another second-line glucose-lowering drug (n = 23,664; 69.33%). After weighting, baseline demographic characteristics were generally comparable between treatment groups. The mean age was slightly lower among SGLT2i initiators compared with other-GLD users (65.47 ± 9.03 vs. 66.06 ± 9.66 years; P < 0.001). The distribution of sex was similar across groups (female: 48.93% vs. 49.66%; P = 0.06). Racial and ethnic composition also did not differ meaningfully, with comparable proportions of Hispanic (13.87% vs. 13.91%), non-Hispanic Black (26.26% vs. 27.03%), and non-Hispanic White patients (42.99% vs. 43.03%; P = 0.0247).

Clinical comorbidities showed modest between-group differences. Prevalence of atrial fibrillation was similar (11.48% vs. 11.13%; P = 0.15), whereas SGLT2i users had slightly higher frequencies of heart failure (15.62% vs. 15.01%; P = 0.0273) and chronic kidney disease (16.82% vs. 16.25%; P = 0.0478). Anemia was marginally less common among SGLT2i initiators (19.44% vs. 20.10%; P = 0.0324). Use of insulin at baseline was slightly lower in the SGLT2i group (33.44% vs. 34.50%; P = 0.004). The proportion of individuals with an AKI diagnosis prior to follow-up was lower among SGLT2i initiators compared with other-GLD users (0.83% vs. 1.30%; P < 0.001). Mean follow-up time for incident AKI also differed between groups, with shorter follow-up observed among SGLT2i users (1.18 ± 2.10 vs. 2.87 ± 2.86 years; P < 0.001).

### Primary effect estimation

For Step 1: primary estimation, we follow a TTE framework. SGLT2i have a trend to reduce the risk but not significant (HR, 0.87, 95%CI [0.75, 1.02]; P=0.087). A pre-defined subgroup analysis reveals that for young population (age < 65), SGLT2i have association with a reduced risk of AKI compared to other second-line GLDs (HR, 0.72, 95%CI [0.57, 0.9]; P=0.004).

### Heterogeneous treatment effects

The estimated risk difference (RD) was negative for AKI (RD: -0.004, 95% CI: -0.006 to 0.001) indicating a potential benefit, though the CIs remain wide. The result was consistent with Cox regression model.

### Interpretable Tree Analysis

We employed single decision tree models to identify patient subgroups with varying SGLT2i effects. The causal interpretable decision tree revealed meaningful heterogeneity in the estimated treatment effects of SGLT2 inhibitor initiation on AKI risk across clinically distinct subgroups. The root split was driven by atrial fibrillation (AF), indicating AF as the strongest effect modifier in the cohort. Among patients *without* AF (N = 34,480), the RD suggested a modest protective association (RD = –0.004). This subgroup further divided on use of anti-Parkinson agents, identifying a clinically coherent pathway: individuals without AF and not using antiparkinsonian agents (N = 28,192) showed a small but statistically robust reduction in AKI risk (RD = –0.004; 95% CI: –0.007 to –0.001). In contrast, those without AF but using anti-Parkinson agents (N = 2,399) exhibited a substantially stronger protective effect (CATE = –0.014; 95% CI: – 0.023 to –0.005), highlighting a distinct high-benefit subgroup.

Among patients *with* AF (N = 3,889), the tree identified anemia as an additional effect modifier. Patients with AF and *no* anemia (N = 2,292) showed a slight increase in AKI risk associated with SGLT2i initiation (RD = 0.007; 95% CI: 0.000 to 0.014), while those with both AF and anemia (N = 1,597) exhibited a near-null effect (RD = 0.001; 95% CI: –0.007 to 0.008).Detailed interpretable tree for AKI was provided in **Figure 4**.

**Figure 3a.**
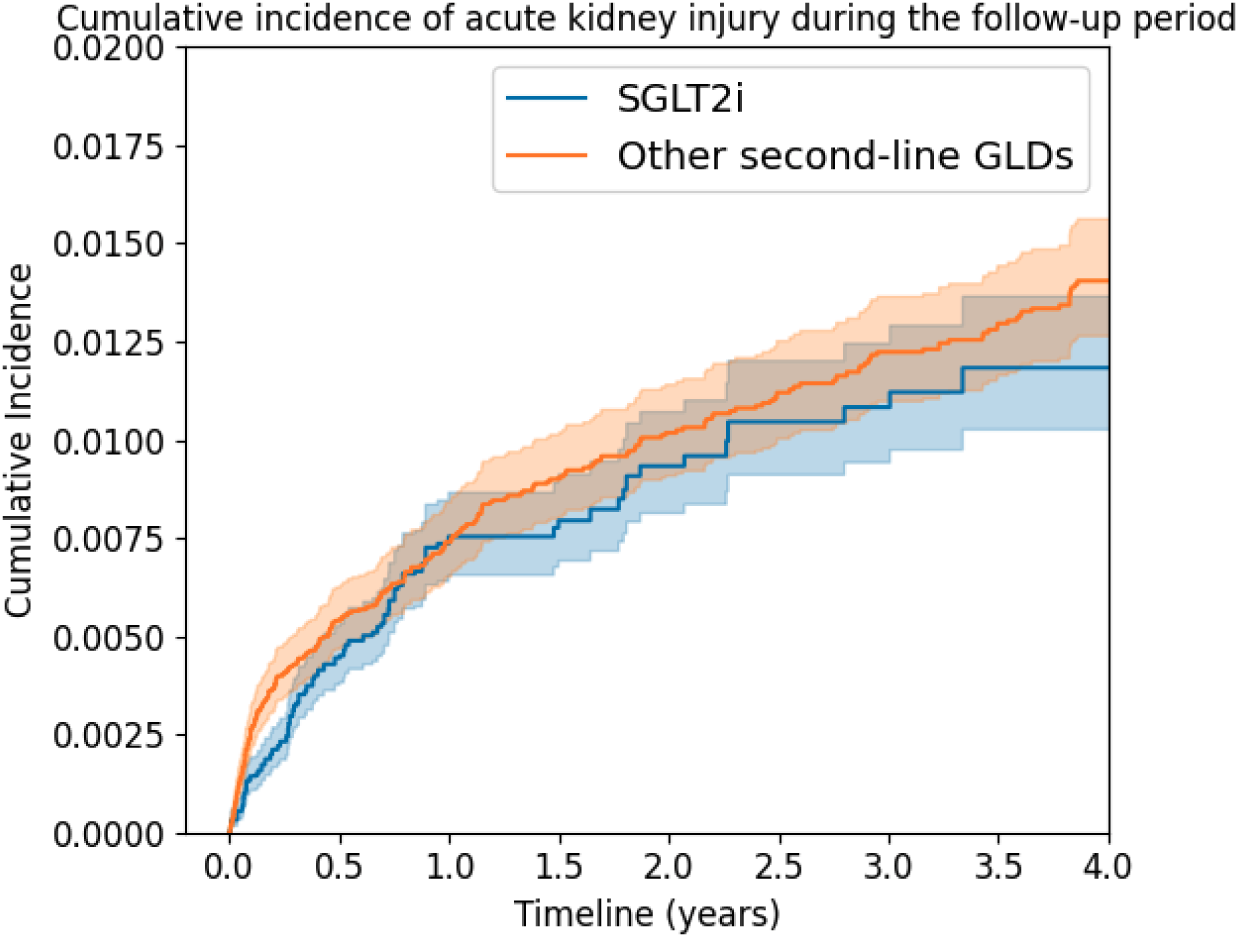
Incidence rate of AKI.

**Figure 3b.**
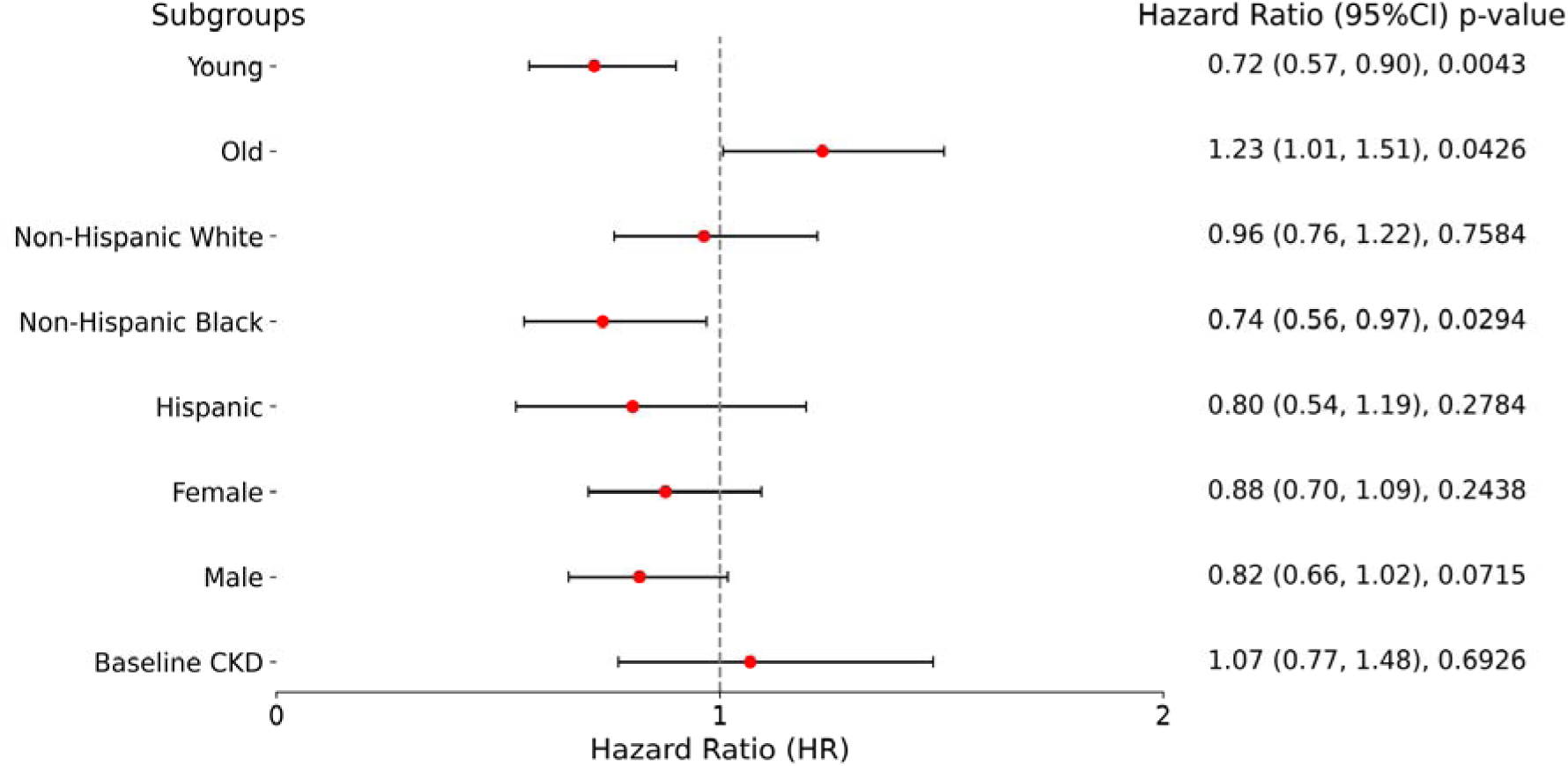
Subgroup analysis.

**Figure 4.**
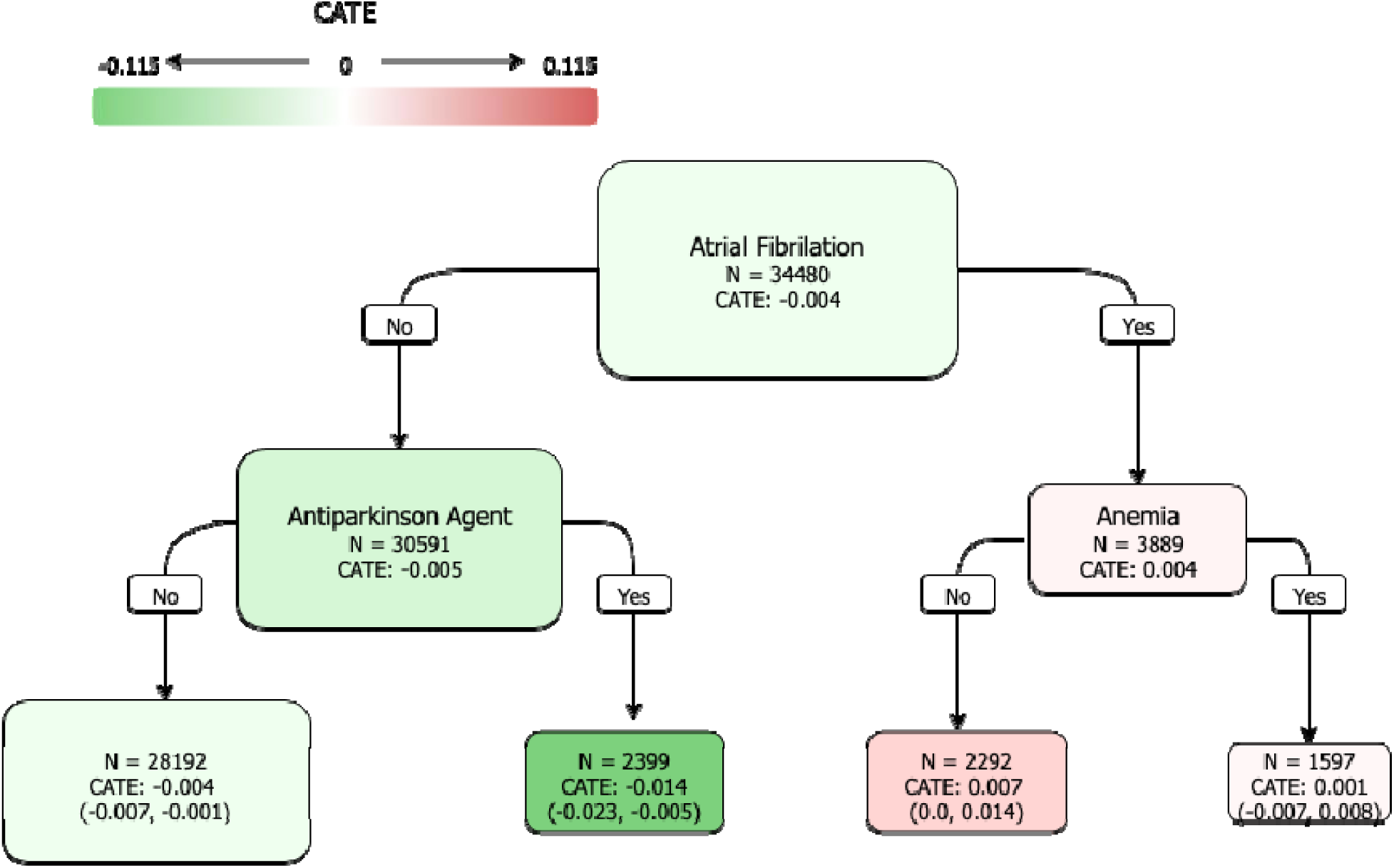
Interpretable decision tree for HTE.

### Causal Structure Discovery

Data-driven causal structure learning using the BOSS algorithm identified a coherent and biologically plausible directed acyclic graph linking demographic factors, comorbidities, medications, kidney function, treatment exposure, and AKI risk (**Figure 5a**). Across the full cohort, eGFR emerged as the most proximal determinant of AKI, with direct incoming edges from chronic kidney disease, anemia, atrial fibrillation, and age, indicating a multicomponent upstream structure of renal vulnerability. Treatment assignment (SGLT2i vs. other GLDs) was influenced by several clinical characteristics, most notably insulin use, heart failure, obesity, and baseline GLP-1RA therapy, highlighting important confounding pathways in real-world prescribing. Notably, treatment assignment did not exhibit a direct edge to AKI in the learned graph; instead, its relationship with AKI appeared to operate predominantly through eGFR, suggesting potential mediation via kidney function.

To further probe effect modification suggested by the HTE analyses, separate causal graphs were learned within subgroups defined by atrial fibrillation, age, and insulin use (**Figure 5b**). In the atrial fibrillation subgroup, heart failure and anemia formed a tightly connected cluster influencing both CKD and eGFR, with eGFR again serving as the principal gateway to AKI. Within the older age subgroup, age showed a stronger direct influence on eGFR, and treatment selection pathways were more heavily shaped by obesity and GLP-1RA use. In the insulin-treated subgroup, insulin, opioid exposure, and GLP-1RA therapy formed a distinct prescribing pathway that shaped variability in treatment assignment, with downstream propagation to eGFR and AKI.

**Figure 5a.**
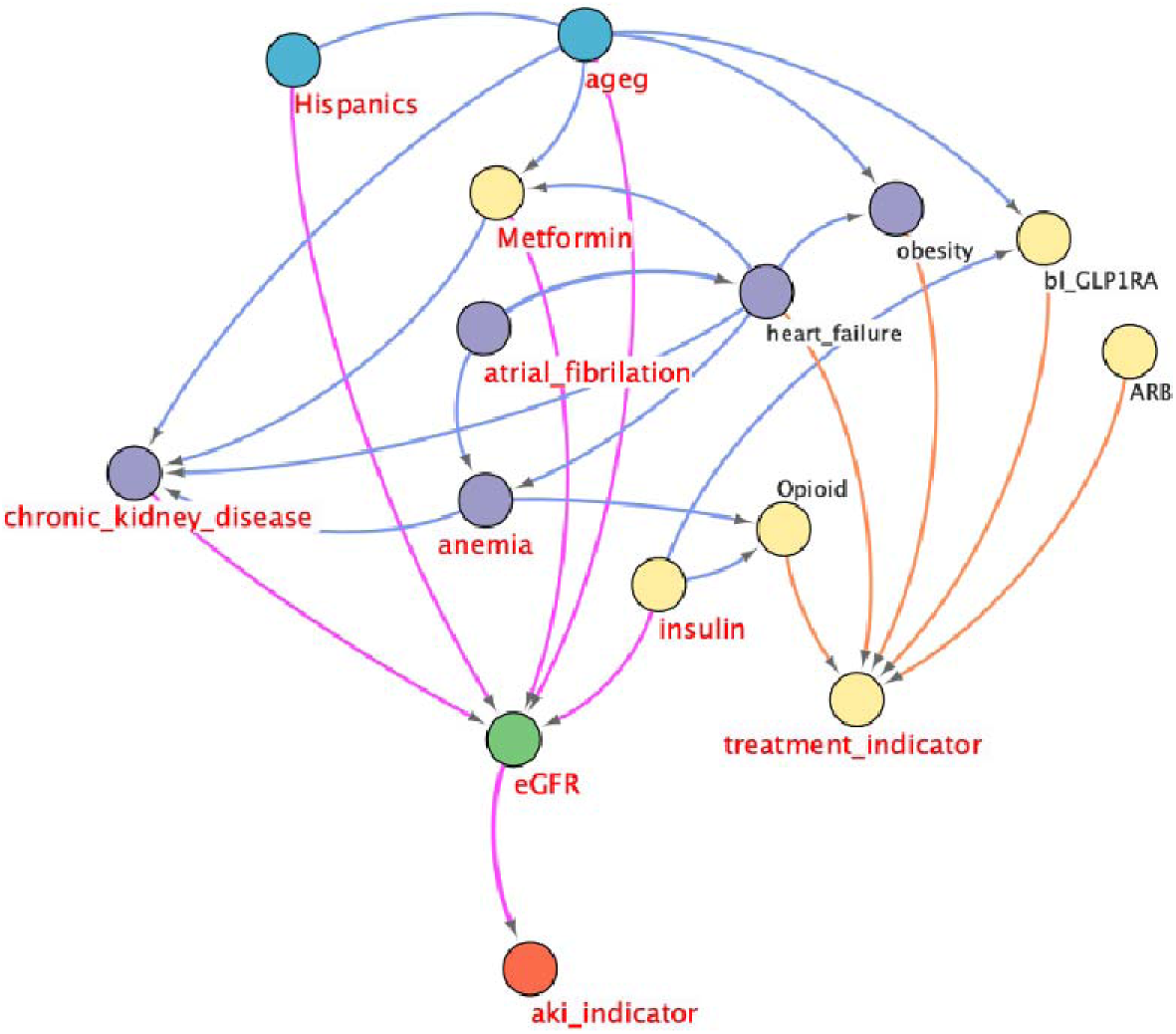
DAG.

**Figure 5b.**
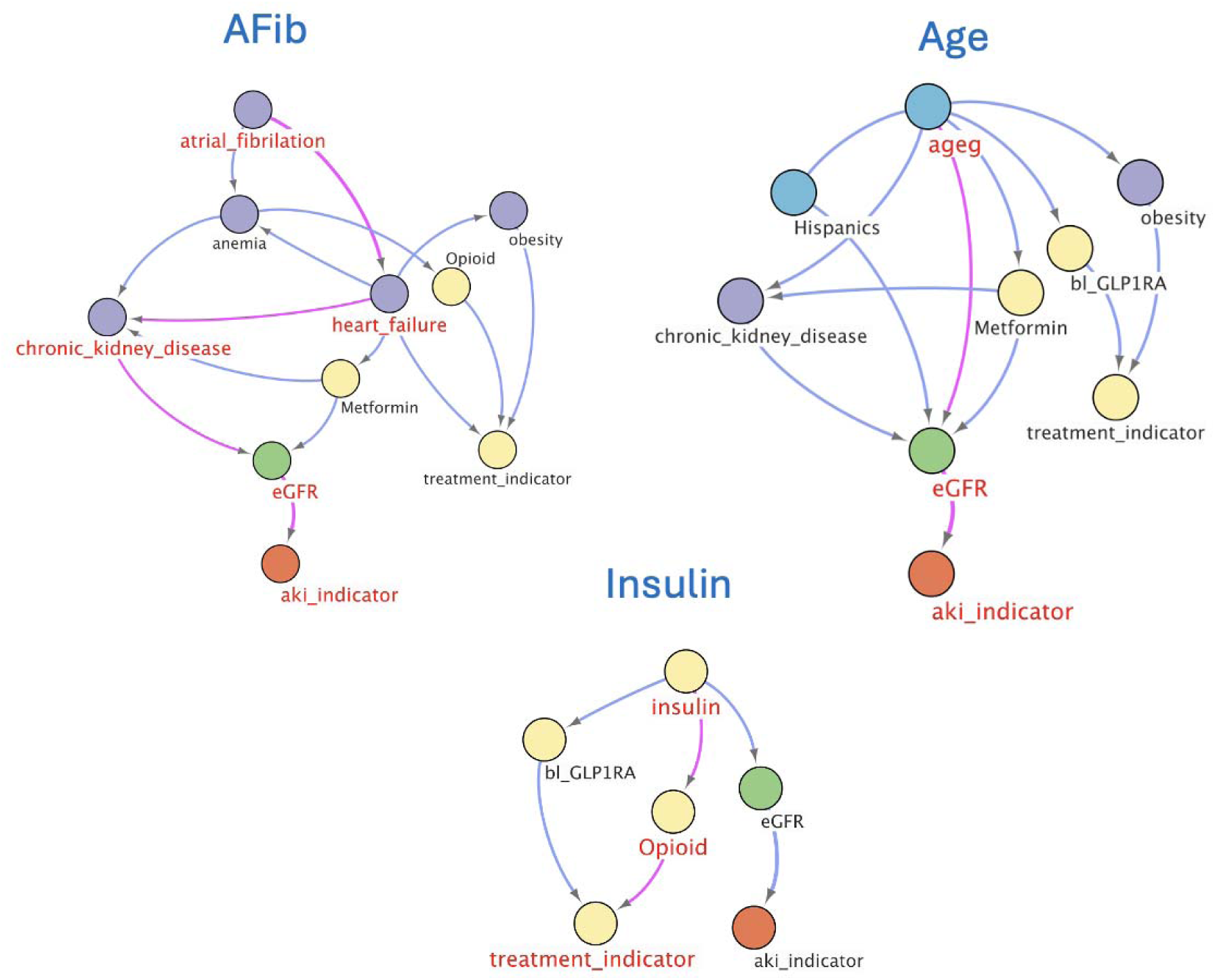
Sub-DAG.

**Figure 6a.**
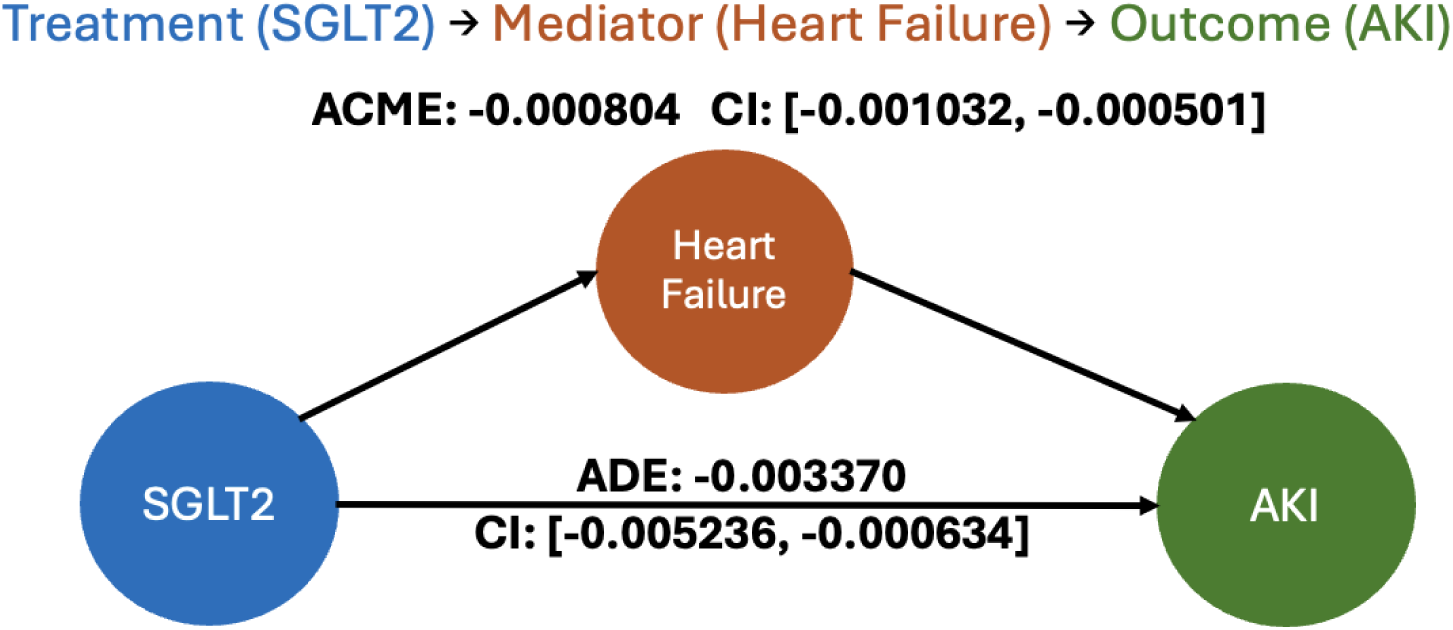
Mediation effect of heart failue.

**Figure 6b.**
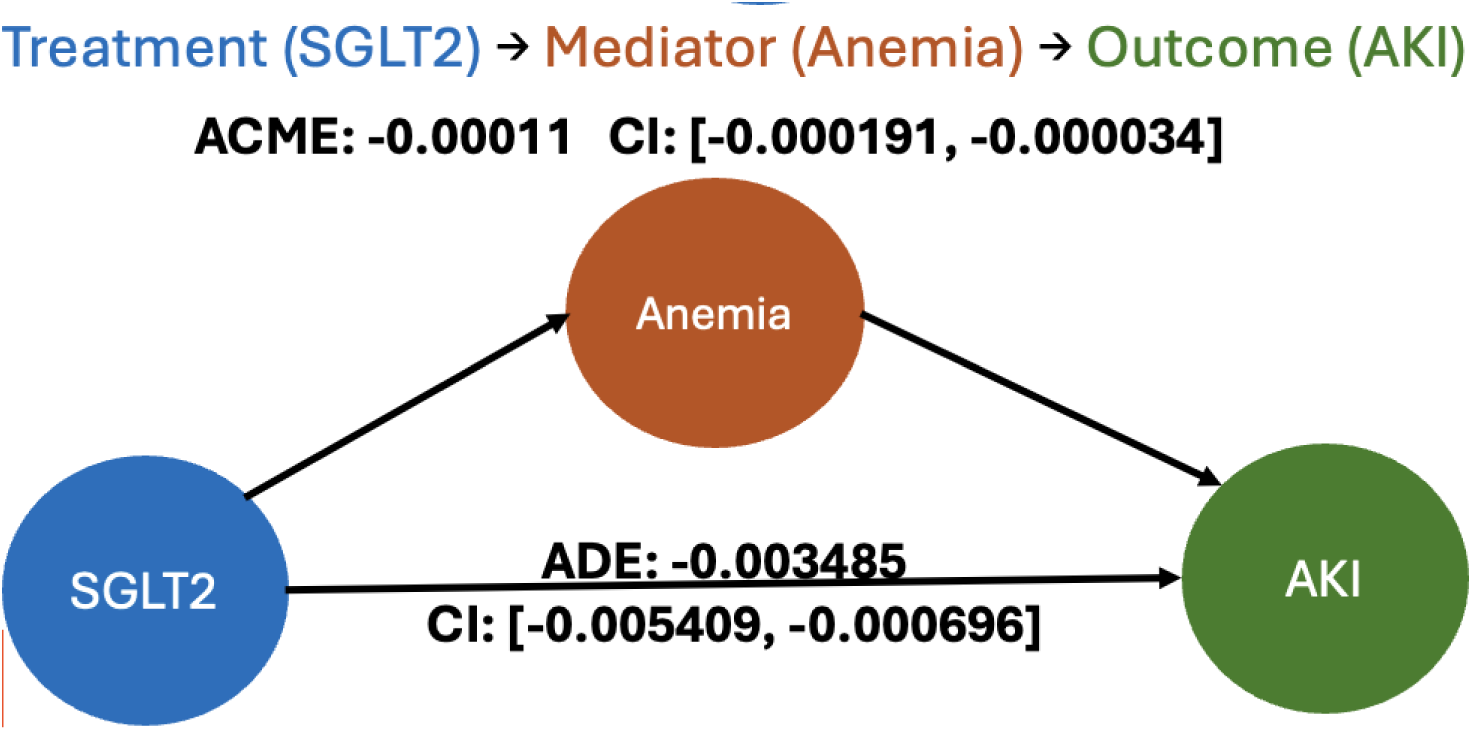
Mediation effect of anemia.

**Figure 6c.**
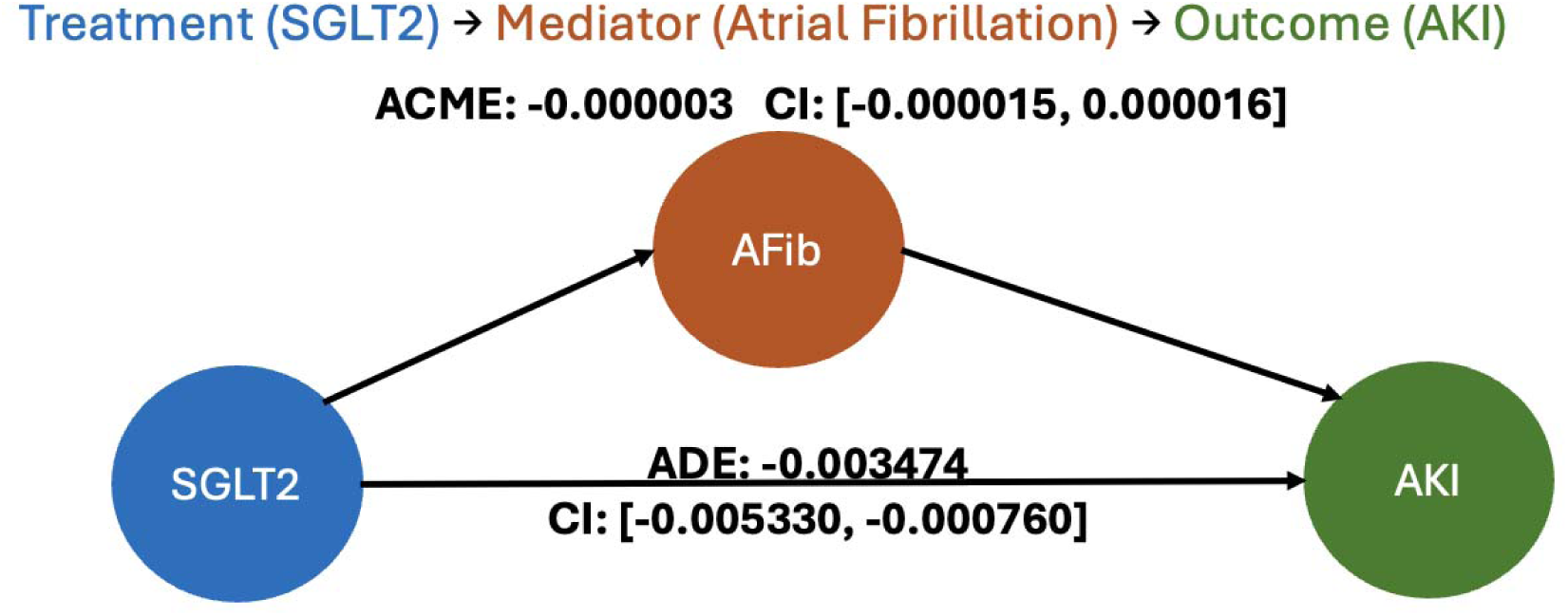
Mediation effect of heart atrial fibrillation.

Collectively, these causal graphs delineate the underlying directional relationships among patient characteristics, treatment choice, kidney function, and AKI. They highlight eGFR as the dominant mediator, identify subgroup-specific confounding structures, and provide mechanistic context for the heterogeneous treatment effects observed in earlier analyses.

### Causal paths decomposition

Using the FACTS framework, we decomposed the total estimated treatment effect into a set of directed causal pathways linking SGLT2i initiation to AKI risk (**Table 2**). Multiple pathways contributed meaningfully to the overall effect; however, two criteria were used to identify the most influential pathway: (1) the magnitude of its contribution to the total effect, and (2) the number of component nodes, as longer pathways may capture clinically important indirect processes that are otherwise hidden in conventional mediation analyses.

**Table 2.** FACTS.

| Path | dp_contribution | acc_contribution |
| --- | --- | --- |
| (AF, HF, CKD, eGFR) | -0.017205 | 0.014849 |
| (AF, anemia) | -0.01559 | 0.013148 |
| (AF, HF, anemia) | -0.00869 | 0.007386 |
| (AF, HF, Metformin, eGFR) | -0.008166 | 0.006883 |
| (AF, HF, CKD) | -0.006463 | 0.005723 |
| (AF, anemia, CKD, eGFR) | -0.003231 | 0.002668 |
| (AF, HF, anemia, CKD, eGFR) | -0.002882 | 0.002552 |
| (AF, HF, Metformin, CKD, eGFR) | -0.002074 | 0.001837 |
| (AF, HF, Metformin) | -0.001157 | 0.000715 |
| (AF, anemia, CKD) | -0.000568 | 0.000503 |
| (AF, HF, Metformin, CKD) | -0.000459 | 0.000406 |
| (AF, HF, anemia, CKD) | -0.000415 | 0.000367 |
| (AF, HF, anemia, Opioid, SGLT2i) | -0.000284 | 0.000251 |
| (AF, anemia, Opioid, SGLT2i) | -0.000175 | 0.000155 |
| (AF, HF, anemia, Opioid) | 0 | 0 |
| (AF, anemia, Opioid) | 0.000437 | -0.000387 |
| (AF, HF, obesity, SGLT2i) | 0.000546 | -0.000483 |
| (AF, HF, obesity) | 0.00107 | -0.000831 |
| (AF, HF, SGLT2i) | 0.006245 | -0.005568 |
| (AF, HF) | 0.152074 | -0.129254 |

AF: atrial fibrillation; CKD: chronic kidney disease; HF: HF;

Among all identified pathways, the sequence atrial fibrillation → heart failure → anemia → chronic kidney disease → eGFR → AKI exhibited both the largest proportional contribution and the highest number of intermediate nodes, indicating a clinically coherent and mechanistically rich pathway. Given its prominence under both criteria, this pathway was selected for further evaluation.

### Mediation analysis

We subsequently conducted causal mediation analysis to quantify the extent to which these intermediate clinical conditions, particularly eGFR and its upstream contributors, mediated the association between SGLT2i use and AKI. This targeted mediation assessment allowed us to isolate the indirect effects operating through the AF–HF–anemia–CKD–eGFR pathway and to evaluate how much of the overall treatment benefit could be attributed to changes along this specific causal chain.

We evaluated the mediating roles of anemia, heart failure, and atrial fibrillation in the association between SGLT2i initiation and AKI. Anemia demonstrated a small but statistically significant indirect effect (ACME = –0.00011; 95% CI: –0.000191 to – 0.000034), with a larger direct effect from treatment to AKI (ADE = –0.003485; 95% CI: –0.005409 to –0.000696). Heart failure showed a stronger mediating contribution (ACME = –0.000804; 95% CI: –0.001032 to –0.000501), accompanied by a consistent direct effect (ADE = –0.003370; 95% CI: –0.005236 to –0.000634). In contrast, atrial fibrillation exhibited a negligible indirect effect (ACME = –0.000003; 95% CI: –0.000015 to 0.000016), indicating minimal mediation through this pathway, while the direct effect remained similar to other models (ADE = –0.003474; 95% CI: –0.005330 to –0.000760). Overall, heart failure and anemia contributed modest but detectable indirect effects, whereas atrial fibrillation did not appear to mediate the treatment–outcome relationship.

## Discussion

In this large, real-world cohort of adults with type 2 diabetes, we used a comprehensive causal learning framework to examine the association between SGLT2 inhibitor initiation and the risk of acute kidney injury. Although the primary analysis suggested a nonsignificant trend toward reduced AKI risk overall, our investigation revealed substantial heterogeneity across clinical subgroups and highlighted important causal pathways underlying this relationship. These findings complement and extend previous observational and randomized studies reporting reduced AKI risk among SGLT2i users.

A major contribution of this study lies in leveraging doubly robust meta-learning and causal interpretable decision trees to uncover granular heterogeneity in treatment effects. Whereas traditional pharmacoepidemiologic studies typically estimate only average associations, our results demonstrate that the reno-protective effects of SGLT2is are not uniform across patient populations. Consistent with emerging interest in treatment effect heterogeneity in cardio-renal-metabolic therapeutics,8,9 our model identified atrial fibrillation as the strongest effect modifier, separating subgroups with distinct risk profiles. Patients without AF, and especially those without Parkinson-related medication use, experienced the clearest protective effect, whereas individuals with AF and concomitant anemia showed attenuated or slightly adverse estimates. These findings emphasize the importance of moving beyond average treatment effects to inform more individualized therapeutic decisions.

Our causal structure discovery analyses further illuminated the underlying mechanisms linking SGLT2i therapy to AKI risk. Across the full cohort, eGFR consistently emerged as the most proximal determinant of AKI, aligning with well-established renal physiology and prior literature on kidney function as the dominant predictor of AKI vulnerability.1,3 The BOSS-derived causal graph also demonstrated that treatment assignment was shaped by multiple clinical variables, such as heart failure, obesity, baseline GLP-1RA use, and insulin therapy, reflecting real-world prescribing complexity and underscoring the need for advanced causal adjustment strategies. Notably, no direct causal edge between SGLT2i treatment and AKI was observed, suggesting that kidney function–related pathways may mediate a substantial portion of the treatment effect.

Using the FACTS framework, we decomposed the treatment effect into multiple directed paths and identified a clinically coherent and mechanistically rich chain: atrial fibrillation → heart failure → anemia → CKD → eGFR → AKI, as the most influential. This aligns with the known interdependence between cardiac dysfunction, hematologic abnormalities, and renal impairment, which together drive AKI susceptibility in high-risk populations. Our subsequent mediation analysis quantified these relationships, demonstrating modest but significant indirect effects through anemia and heart failure. Such findings provide empirical evidence supporting the hypothesis that SGLT2i-associated renal benefits may operate, at least partly, through reduction of intermediate cardio-renal stressors rather than solely direct renal mechanisms.

Taken together, these results underscore that the SGLT2i and AKI association is multifactorial, heterogeneous, and largely mediated by intermediate clinical states. This represents a conceptual shift from viewing SGLT2is as uniformly nephroprotective to understanding their benefits as conditional upon patient-specific cardiovascular and renal profiles.

Several limitations warrant consideration. First, despite the use of advanced causal inference techniques, residual confounding cannot be fully excluded, particularly for unmeasured clinical factors such as provider prescribing preference or subclinical kidney injury. Second, ICD-based AKI identification may underestimate event incidence, though algorithmic definitions are widely used in comparative effectiveness research.6 Third, mediation analyses remain sensitive to model specification and sequential ignorability assumptions; however, the consistency across anemia- and heart failure mediated pathways supports robustness. Finally, follow-up time was shorter among SGLT2i users, which may have influenced event detection, though weighting and model-based approaches mitigate most imbalances.

Despite these limitations, our study demonstrates the value of integrating causal machine learning, interpretable subgroup discovery, and path decomposition to enhance mechanistic understanding in pharmacoepidemiology. This framework allows the field to advance beyond average associations toward more individualized, mechanism-informed clinical insights.

## Conclusion

Using a comprehensive causal learning strategy applied to a large, diverse real-world dataset, we found that SGLT2i initiation was associated with a modest but heterogeneous reduction in AKI risk. The renoprotective effect varied across patient subgroups, with distinct benefits observed in individuals without atrial fibrillation and without Parkinson-related medication use. Causal structure discovery and path decomposition highlighted the central role of kidney function and upstream cardio-hematologic conditions in mediating treatment effects, while mediation analyses confirmed that heart failure and anemia contributed small but significant portions of the overall effect.

These findings suggest that the impact of SGLT2is on AKI is not uniform but is shaped by interconnected cardiovascular, hematologic, and renal pathways. Integrating causal ML–based heterogeneity and mechanistic analyses provides a deeper understanding of drug–outcome relationships and offers a foundation for more personalized therapeutic strategies. Future studies should validate these findings in external populations and explore whether tailoring SGLT2i use according to mechanistic risk profiles improves renal outcomes.

## Author Contributions

Dr. J. Guo is the guarantor of this work and, as such, had full access to all the data in the study and takes responsibility for the integrity of the data and the accuracy of the data analysis.

Concept and design: J. Guo, and J. Bian.

Acquisition, analysis, or interpretation of data: All authors. Drafting of the manuscript: H. Dai, Y. Lee

Critical review of the manuscript for important intellectual content: All authors. Statistical analysis: H. Dai and Y. Lee

Obtained funding: J. Guo. Supervision: J. Guo and J. Bian.

## Data Availability Statement

Data set Available through OneFlorida+ Clinical Research Network (email)

